# Spinal cord stimulation modulates post-synaptic inhibition improving neuromotor control of arm movement in people with chronic hemiparesis post-stroke

**DOI:** 10.1101/2025.09.23.25336157

**Authors:** Luigi Borda, Nikhil Verma, Erynn Sorensen, Erick Carranza, Roberto de Freitas, Scott Ensel, Amy Boos, Peter C. Gerszten, Daryl P. Fields, George F. Wittenberg, Lee E. Fisher, Elvira Pirodini, Marco Capogrosso, Douglas J. Weber

## Abstract

Epidural spinal cord stimulation has been shown to be a promising neurotechnology to improve upper limb function in people affected by stroke. It is well established that SCS targeting the dorsal root entry zone increases excitatory drive to α-motoneurons via the monosynaptic reflex pathway. However, the effects of SCS on inhibitory neural pathways remain unexplored. We hypothesized that SCS improves the neuromotor control of arm movement by strengthening both excitatory and inhibitory circuit function. We show in three individuals with post-stroke motor symptoms that SCS enhances postsynaptic Ia reciprocal inhibition. These changes correlate with enhanced muscle coordination and arm kinematics resulting in smoother and faster trajectories. Our study provides further insights into the neural targets of spinal cord stimulation and the use of this technology to treat post-stroke motor deficits.

## INTRODUCTION

Voluntary movements require functional communication of motor commands from the brain to motor circuits located in the spinal cord^1,2^. To generate smooth and accurate movement, supraspinal commands regulate activation of multiple muscle groups throughout movement initiation and execution^3–5^. After stroke, damage to the corticospinal tract leads to motor impairments characterized by muscle weakness, excessive agonist/antagonist co-activation and intrusion of flexor and extensor synergies^6–9^. At present, activity-based training is the most common therapy for alleviating post-stroke impairments ^10,11^. Through repetitive task training, stroke survivors exploit and strengthen intact motor pathways thus promoting restorative plasticity^12^. Thus, the ability to produce voluntary movement is crucial to engage the neurophysiological mechanisms that drive recovery of motor function. This has prompted the development of neurotechnologies aimed at assisting rehabilitation programs, particularly in patients with severe impairments that have limited ability to participate in therapy^13^.

Our group has shown recently that tonic stimulation of primary afferent neurons using spinal cord stimulation (SCS) represents a promising avenue for promoting the recovery of motor functions post-stroke^14^. Traditionally, it has been thought that SCS exploits the monosynaptic reflex pathway to evoke a volley of excitatory inputs to the α-motoneurons^15–19^ making them more responsive to weak supraspinal motor commands hence facilitating movement execution^20^. However, the influence of excitatory inputs from the sensory neurons may not be limited to monosynaptic connections. Polysynaptic pathways involving excitatory and inhibitory interneurons may also be engaged by stimulation and therefore play a critical role in the recovery of motor functions. Decades of research has indeed revealed that Ia afferents share both a monosynaptic connection with the α-motoneurons innervating the homonymous muscle and a disynaptic connection via the Ia inhibitory interneuron with the α-motoneurons innervating the heteronymous muscles^21–26^. Moreover, computer simulations have shown that the enhancement of this inhibitory pathway is critical in the generation of smooth and effective control of locomotor function after a neural lesion^27^.

Here, we hypothesized that the immediate improvements mediated by SCS are due to a concurrent enhancement of excitatory and inhibitory circuit function. Specifically, we investigated whether postsynaptic Ia reciprocal inhibition^23,26^ would be increased during SCS. To this end, we performed a battery of experiments in three individuals with chronic stroke and found that SCS improved the ability of participants to reduce antagonist muscle activity. We also show that SCS is directly involved in the modulation of the antagonist motor pool excitability with timing consistent with the short-latency inhibition mediated by the Ia inhibitory interneuron. Taken together our results suggest that the excitatory inputs evoked by SCS also engage inhibitory interneurons to reduce agonist/antagonist coactivation thus improving the neuromotor control of arm movements.

## RESULTS

### Study participants and spinal cord stimulation leads placement

In this manuscript, we report the results from three participants in a clinical study (NCT04512690, see methods) aiming to investigate the effects of epidural spinal cord stimulation for treating upper extremity motor deficits after stroke. All participants were 4-10 years out from their stroke, placing them well within the chronic phase of their stroke recovery and affected by upper-limb hemiparesis. The baseline demographics at the time of enrollment in the study are reported in Table 1 and Figure 1a,b. The pre-study Fugl-Meyer (FM, see methods) scores were 23 (SCS04), 34 (SCS07) and 32 (SCS08) out of 66 for the motor assessments; the FM sensory scores, instead, were 3 (SCS04), 11 (SCS07) and 12 (SCS08) out of 12 (Table 1; Figure 1b).

**Figure 1.**
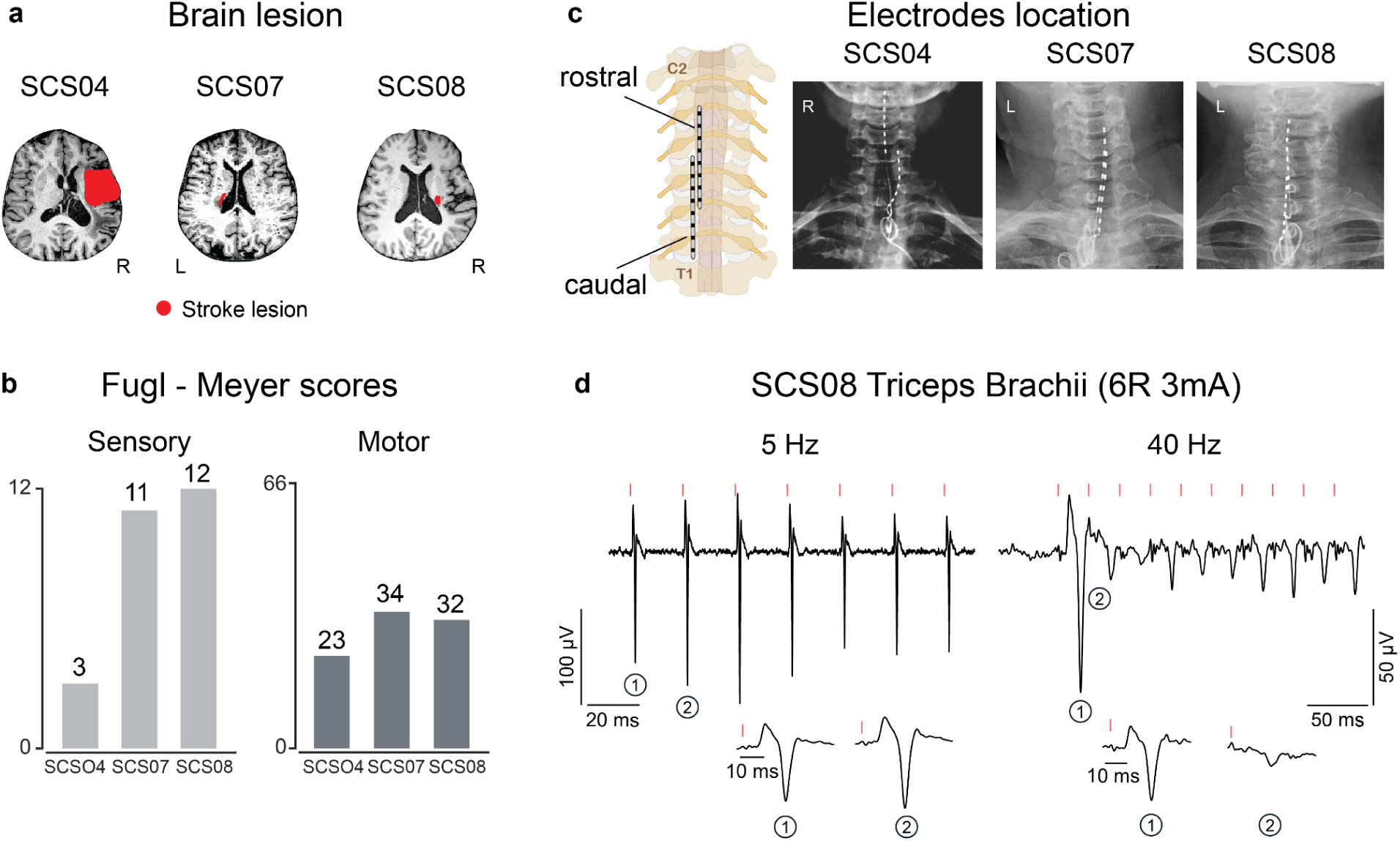
Study participants and SCS leads placement: **a**) MRI scan of each participant with the segmented lesion indicated in red. **b**) Fugl-Meyer scores for both sensory and motor assessments for all the participants. **c**) X-rays of the three participants showing the location of the contacts for the rostral and caudal lead. **d**) Example of frequency dependent depression^18,29^ (SCS08); we delivered tonic SCS at 5Hz and 40Hz through the sixth contact of the rostral lead while recording electromyographic (EMG) activity from the triceps brachii. We observed a clear decrease in the peak-to-peak amplitude of the responses evoked during 40 Hz stimulation indicating presynaptic recruitment of the motoneurons through the proprioceptive fibers.

**Table 1.** Demographics.

| ID | Sex | Stroke type | Lesion Area | Time since stroke | FM motor | FM sensory |
| --- | --- | --- | --- | --- | --- | --- |
| SCS04 | M | Ischemic | Right cortical hemisphere | 5 years | 23 | 3 |
| SCS07 | F | Ischemic | Left cortical hemisphere | 10 years | 34 | 11 |
| SCS08 | F | Ischemic | Right cortical hemisphere | 4 years | 32 | 12 |

Two 8-contact SCS leads were implanted percutaneously in the epidural space overlying the dorsal root entry zone of the cervical spinal cord. To target the upper-limb muscle motor pools, the leads were placed spanning C3-T1 spinal segments as we described previously^14^ (Figure 1c). The percutaneous leads were implanted for 29 days and then removed. During the implant period, the individuals participated in testing sessions five times per week, each lasting approximately 4 h.

It is well established that subthreshold SCS does not produce direct activation of motoneurons, but instead facilitates voluntary recruitment of motoneurons by driving synaptic inputs to the spinal motoneurons and interneurons^14,18,19^. To confirm the correct placement of the leads across spinal segments and ensure the trans-synaptic activation of motoneurons through the sensory fibers, we verified that the reflex-mediated responses exhibited frequency-dependent depression^18,29^. Figure 1d shows examples of the compound muscle action potentials (CMAPs) evoked in the triceps brachii muscle during SCS at 5 and 40 Hz. During 5 Hz stimulation, each stimulation pulse evoked a large CMAP in the triceps whereas during 40 Hz stimulation, the first stimulation pulse evoked a large amplitude CMAP and subsequent stimulation pulses evoked smaller CMAPs. This rate-dependent depression of evoked responses therefore confirmed the pre-synaptic recruitment of α-motoneurons mediated by the Ia sensory afferents.

### Improvements in reaching kinematics are associated with reduced antagonist activity during spinal cord stimulation

To assess if SCS engages reciprocal inhibitory pathways driving motor improvements, we first studied how the arm kinematics and muscle activity changed when participants performed a planar reaching task with and without SCS. Specifically, we asked the participants to reach from a central location towards three targets using a robotic exoskeleton platform (KINARM, see methods) (Figure 2a, top panels). Based on the recruitment curves (see methods), we identified a subset of stimulation electrodes and amplitudes that mainly recruited the elbow extensor muscles (i.e., agonist muscles) to assist the reaching movement (Figure 2a bottom left panel and Extended Data Figure 1a top and bottom left panels). We then analyzed the arm kinematics and compared the effects of the stimulation across three simple and well-established metrics^30^ (see methods): time to reach the target (RT), number of peaks in the elbow speed profile (SP) which indicates the smoothness of the movement, and path efficiency (PE) which indicates the straightness of the arm trajectories (Figure 2b). All participants showed immediate improvements when stimulation was on. For SCS04, who had the most severe post-stroke deficits, movement was both faster and smoother compared to the stimulation off condition (Figure 2b top and bottom left panels; −57.8% RT, −42.7% SP). Similarly, arm movements for SCS07 were both smoother and faster (Figure 2b bottom middle panel and Extended Data Figure 1b top panels; −10.2% RT, −27.9% SP) while movement for SCS08 was both faster and with straighter reaching trajectories (Figure 2b bottom right panel and Extended Data Figure 1b bottom panels; −19.1% RT, −58.4% PE). Because participants could always feel paresthesias^31^ when the stimulation was active, we performed sham stimulation trials to control for motivational bias. In the sham trials, we selected stimulation electrodes and amplitudes that preferentially recruited muscle groups antagonistic to the reaching movement (i.e., elbow flexors, Figure 2a bottom right panel and Extended Data Figure 1a top and bottom right panels). We expected that the sham stimulation would have either no effect or a negative effect on the reaching movement.

**Figure 2.**
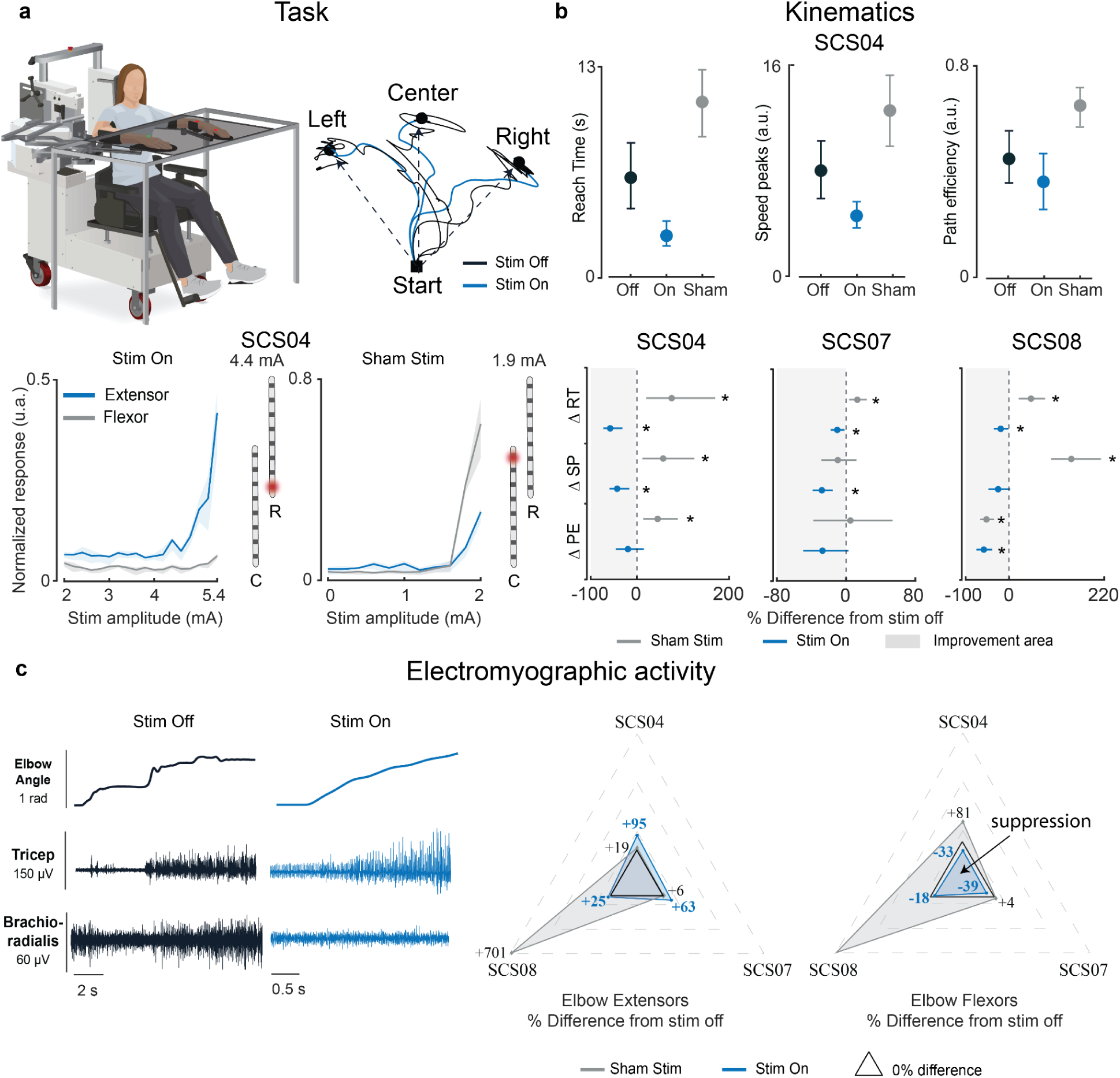
Planar reaching kinematics and muscle activity: **a**, Top left: Participants performed planar reaching movements using a KINARM exoskeleton to three targets as fast and accurately as they could using the paretic arm. Top right: end point trajectories for SCS04 when the stimulation was on (blue) and off (black). Bottom panels show examples of recruitment curves for the contacts 8R and 1C and the normalized evoked responses for triceps brachii (blue, elbow extensor) and brachioradialis (gray, elbow flexor). Thick lines represent the mean values while the shaded areas are the 95% confidence interval (CI) computed with bootstrap N=10,000. **b**, Top panels: mean values of the kinematics metrics used to assess the effect of SCS (On) and sham stimulation with respect to stimulation off for SCS04. Bottom panels: percent change with respect to stimulation off for the two stimulation protocols (On vs Off and Sham vs Off) across all the kinematics metrics (reaching time (RT); speed peaks (SP); path efficiency (PE)). **c**, Left: SCS04 example of elbow angle and electromyographic (EMG) activity for the Triceps Brachii and Brachioradialis while reaching the center target without stimulation (black) and with stimulation (blue). Right: spider plots showing the percent change with respect to stimulation off of the mean EMG activity for elbow extensors (SCS04, SCS07 and SCS08 Triceps Brachii) and elbow flexors (SCS04, SCS07 Brachioradialis and SCS08 Biceps Brachii). Significance tests for all comparisons are reported in Extended Data Figure 1c. The middle circle on plots indicates the mean value of the measurements. All error bars indicate the 95% confidence interval of the mean kinematics metric across repetitions, computed with bootstrap N=10,000. The single asterisk indicates statistical significance and rejection of the null hypothesis of no difference with a 95% CI.

For SCS04, the performance with sham stimulation worsened dramatically in all metrics (Figure 2b top and bottom left panels; +75.5% RT, +56.9% SP, +44.9% PE compared to stimulation off). For SCS07 and SCS08 instead, the sham stimulation either worsened the arm kinematics compared to the stimulation off (SCS07, +12.9% RT; SCS08, +49.4% RT, +141,3% SP) or had no effect in the majority of the kinematics measures (SCS08, - 52.2% PE). Taken together, these results suggest that stimulation of the sensory afferents that recruit the agonist motor pool facilitates movement execution. Conversely, sham stimulation had either no effects or worsened motor control.

Next, we examined how the muscle activity changed when SCS was active and if those changes could explain the differences observed in the kinematics. Specifically, we analyzed the changes in the electromyographic (EMG) activity of elbow extensors and flexors when targeted by stimulation. We expected that stimulation of Ia afferent neurons would provide excitatory input via monosynaptic connections to homonymous muscles (triceps brachii) while concurrently recruiting inhibitory interneurons projecting to motoneurons innervating heteronymous muscles (biceps brachii and brachioradialis). Our results show indeed that SCS boosted the action of the elbow extensors, resulting in increased EMG activity compared to stimulation off, (Figure 2c and Extended Data Figure 1c; SCS04 +95%, SCS07 +63%) and reduced EMG activity in the elbow flexors (SCS04 −33%, SCS07 −39%, SCS08 −18%), supporting our hypothesis that SCS may engage reciprocal inhibitory pathways. Furthermore, we found that these results occurred only when tonic SCS was targeting the agonist motor pool whereas the sham stimulation failed to suppress EMG in the flexor muscles. With sham stimulation, participants showed an increase in both elbow flexor (SCS04, +81%, SCS08 +429%) and elbow extensor EMG (SCS04 +19%, SCS08 +701%). Thus, this suboptimal stimulation increased co-activation of elbow extensors and flexors, resulting in motor performance that was similar or worse than without stimulation.

In summary, SCS targeting the agonist motor pool enhanced inhibitory commands which suppressed the activity of muscles opposing the direction of movement. This effect operates synergistically with the facilitatory effects of SCS on the agonist muscles. Taken together these results support our hypothesis of concurrent SCS-mediated inhibitory effects that improve muscle recruitment and motor control.

### Tonic spinal cord stimulation reduces the excitability of the antagonist motor pool

Our previous findings demonstrate a reduction in elbow flexors EMG when SCS was targeting triceps brachii (TRI) to enhance reaching movements. Decades of research show that the disynaptic connection of the Ia sensory afferents can produce short latency inhibition of the heteronymous muscles via Ia inhibitory interneurons^23^ (Figure 3a, top left panel). Since SCS targeting the dorsal roots produces direct activation of Ia afferents, we reasoned that doing so would engage this inhibitory circuit to suppress recruitment of motoneurons that innervate antagonistic muscles. To test this hypothesis, we measured the effects of SCS on antagonist muscle recruitment during electrically evoked posterior-root muscle (PRM) reflexes. The amplitude of the PRM-reflex is used to measure changes in the excitability of the motor pool^32^. PRM reflexes were evoked in the biceps brachii (BIC) by delivering a 2 Hz train of biphasic and charge-balanced pulses on one SCS electrode targeting the BIC motor pool (Figure 3a, bottom right panel). The average peak-to-peak amplitude of the compound muscle action potential (CMAP) evoked by the PRM reflex was measured (see methods) and compared across three conditions: stim off, stim on, and sham (Figure 3b). The PRM reflex amplitude during the stim off condition served as the baseline. During stim on trials, we applied a 40 Hz train of tonic SCS pulses at sub-motor threshold amplitude to an electrode targeting the TRI motor pool. Tonic SCS of the TRI was delivered concurrently with PRM stimulation of the BIC, interleaved with a 1 ms delay to account for the expected short-latency reciprocal inhibition^23^ (Figure 3a, top right panel). Similarly, in the sham condition, a 40 Hz train of sub-motor threshold SCS pulses was applied to an *off-target* electrode (i.e.one not targeting the TRI) while PRM reflexes were measured in the BIC. During testing, participants were seated with their paretic arm supported against gravity by the KINARM exoskeleton. A member of the study group held the elbow extended to prevent participants from actively producing any muscle contraction, minimizing supraspinal influences on spinal reflex modulation (Figure 3a, bottom left panel). Figure 3b (top row) shows examples of CMAP waveforms for PRM reflexes measured during the stim off and on conditions, which clearly show a consistent reduction in the CMAP amplitude during stimulation across all three participants. Indeed, the amplitude of the PRM-reflexes evoked in the BIC was reduced by more than 40% with stim on (Figure 3b bottom panels; SCS04 −41%, SCS07 −48%, SCS08 −40%). To further confirm that the effects could be attributed to reciprocal inhibition, we repeated the experimental protocol under a sham condition where tonic SCS was delivered via a caudal electrode that facilitated recruitment of forearm and hand muscles. In this scenario, the average CMAP amplitudes were similar to those evoked with the stimulation off, confirming the absence of an inhibitory effect on the PRM reflexes (Figure 3b bottom panels, sham condition). These results support our hypothesis that tonic SCS of the sensory afferents can recruit spinal inhibitory circuits, which could then be leveraged by remaining supraspinal connections resulting in better motor control.

**Figure 3.**
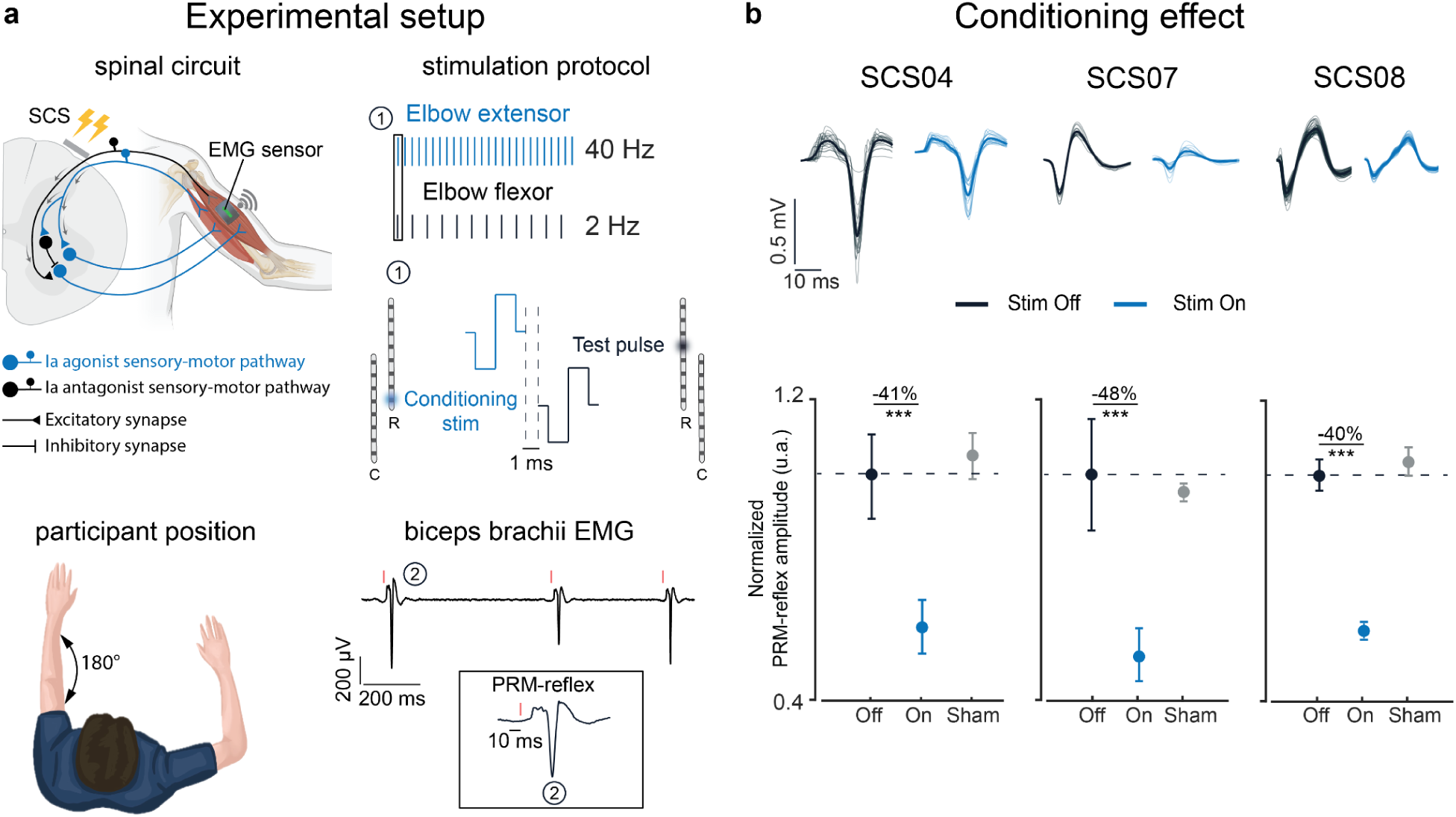
SCS effects on the spinal circuit: **a**, Top left: simplified schematic of the spinal circuit involved in the stimulation protocol. The circuit includes antagonist sensory afferents recruiting α-motoneurons innervating Biceps Brachii (black); agonist sensory afferents recruiting both α-motoneurons innervating the homonymous muscle (blue) (i.e., Triceps Brachii (TRI)) and Ia inhibitory interneurons innervating α-motoneurons of the heteronymous muscle (black) (i.e., Biceps Brachii (BIC)).Top right: example of the stimulation protocol applied to SCS04. Each vertical line is representative of biphasic and charge-balanced pulses concurrently delivered at 40 Hz (conditioning train, agonist) and 2 Hz (test pulses, antagonist). The inset block 1 shows how the test pulse and conditioning train were interleaved with 1 ms delay. The test pulse was delivered through contact 4R whereas the conditioning train was delivered through contact 8R to recruit BIC and TRI sensory afferents, respectively. Bottom left: example of participant with the left paretic arm resting in a fully extended posture. Bottom right: example of the EMG recorded from the Biceps Brachii of SCS04 while delivering only test pulses at 2 Hz through contact 4R. The inset block 2 shows an example of the evoked PRM-reflex recorded in the BIC. **b**, Top: example of the modulation of the PRM-reflexes recorded in the BIC with (blue, stimulation On) and without conditioning stimulation (black, baseline PRM-reflexes when conditioning stimulation was Off). Bottom: quantification of the normalized mean peak-to-peak amplitude of the evoked PRM-reflexes recorded in the BIC for all the participants across the stimulation conditions (Off: 2 Hz test pulses delivered through the contact targeting BIC; On: 2 Hz test pulses targeting BIC interleaved with tonic SCS at 40 Hz targeting TRI; Sham: test pulses at 2 Hz targeting BIC interleaved with tonic SCS at 40 Hz targeting hand/forearm muscles). The circle marker on the plots indicates the mean value of the measurements. All responses were normalized to the mean peak-to-peak amplitude measured in the Off condition. All error bars indicate the 95% confidence interval of the mean normalized peak-to-peak amplitude, computed with bootstrap N=10,000. Statistical significance was assessed with two-tail bootstrapping (N=10,000): p<0.05 (*), p<0.01 (**), p<0.001(***).

### Spinal cord stimulation potentiates central inhibition

After confirming that SCS can engage and modulate the activity of inhibitory pathways in the spinal cord, we tested whether this modulation changes during active movement. Balaguer et al^33^. have indeed shown that even after a neural lesion like stroke or spinal cord injury, residual excitatory and inhibitory supraspinal inputs can modulate the motoneurons’ membrane potential to sculpt voluntary motor output in the presence of SCS. We thus hypothesized that the inhibitory effects produced by the stimulation would be further enhanced during active movement to silence unwanted antagonist activity. To test this hypothesis, we measured PRM-reflex modulation during active elbow extension movement. Participants performed a point-to-point planar reaching task while seated with their arm supported by the KINARM exoskeleton (Figure 4a top panels). We employed the same stimulation paradigm as was used in the previous resting condition (Figure 3a). A 2 Hz train of suprathreshold stimulation pulses was applied at the SCS electrode targeting elbow flexors muscles to evoke PRM-reflexes in the BIC during movement (Figure 4a bottom panels). We then first compared the amount of reflex modulation against the peak-to-peak amplitude of the PRM-reflexes measured at rest with no conditioning stimulation applied (i.e., baseline). We found that during movement, without conditioning stimulation applied, SCS04 who was the participant with the most severe motor impairments (FM = 23) had a 60% reduction in the peak-to-peak amplitude of the PRM reflexes compared to their amplitude at rest. Meanwhile, the less impaired participants, SCS07 (FM = 34) and SCS08 (FM = 32), showed a reduction of 67% and 85%, respectively (Figure 4b, gray bars). The remaining ability to suppress evoked unwanted activity in antagonist muscles during movement was therefore loosely correlated with the level of motor impairment with the most severe participant which showed less reduction compared to the others.

**Figure 4.**
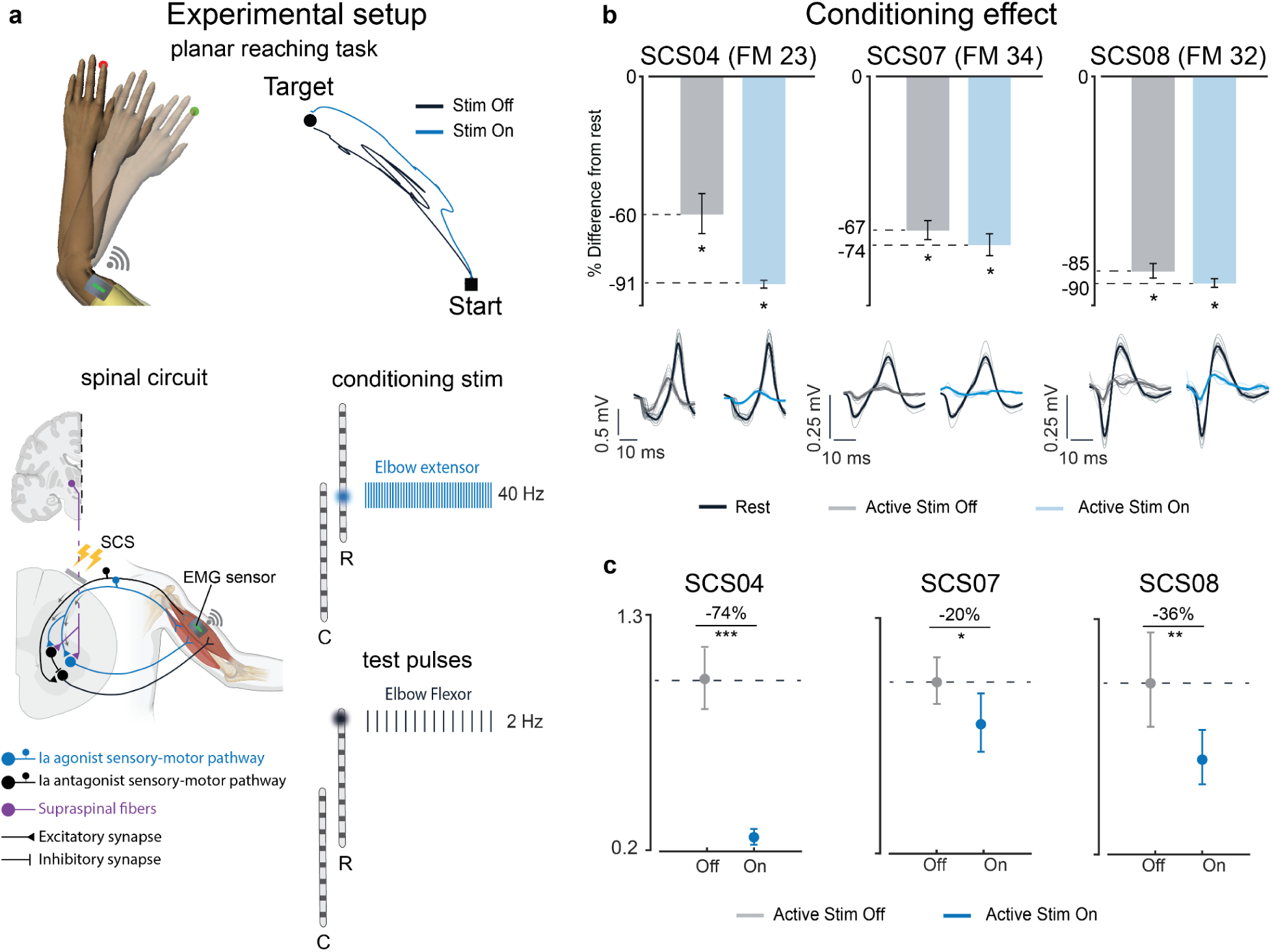
SCS potentiates central inhibition: **a**, Top left: example of the planar reaching task. Participants were asked to make repeated reaching movements to a fixed target. Top right: end point trajectories for SCS04 when the conditioning train was Off (black) and On (blue). Bottom left: simplified schematic of the spinal and supraspinal connections involved. The circuit includes antagonist sensory afferents recruiting α-motoneurons innervating Biceps Brachii (black); agonist sensory afferents recruiting both α-motoneurons innervating the homonymous muscle (blue) (i.e., Triceps Brachii (TRI)) and Ia inhibitory interneurons innervating α-motoneurons of the heteronymous muscle (black) (i.e., Biceps Brachii (BIC)). Corticospinal fibers (violet) recruiting both the Ia inhibitory interneuron (black) and the α-motoneuron innervating Triceps Brachii (blue). Bottom right: example of the stimulation protocol applied during the reaching task. Each vertical line is representative of biphasic and charge-balanced pulses. Test pulses were delivered at 2 Hz through SCS leads contact targeting elbow flexors. Concurrently, conditioning stimulation trains were delivered at 40 Hz through the contact targeting elbow extensors. Test pulses and conditioning trains were interleaved with 1 ms delay. **b**, Quantification of percent change with respect to the peak-to-peak amplitude of the responses evoked in the BIC at rest (baseline) for all conditions during movement (stim off vs baseline in gray; stim on vs baseline in blue). Bar plots indicate the mean percent difference. Error bars indicate the 95% confidence interval of the mean percent difference computed with bootstrap N=10,000. The single asterisk indicates statistical significance and rejection of the null hypothesis of no difference with a 95% CI. Bottom: example of the evoked PRM-reflexes recorded in the BIC during rest (black) and active movement with (blue) and without (gray) condition stimulation. **c**, Quantification of the mean normalized peak-to-peak amplitude of the evoked PRM-reflexes recorded in the BIC for all the participants in all the conditions (Off: only test pulses delivered at 2 Hz through the contact targeting BIC; On: test pulses at 2 Hz targeting BIC interleaved with tonic SCS at 40 Hz targeting TRI). The circle marker on the plots indicates the mean value. All responses were normalized to the mean peak-to-peak amplitude measured in the off condition. The error bars indicate the 95% confidence interval of the mean peak-to-peak amplitude, computed with bootstrap. Statistical significance was assessed with two-tail bootstrapping (N=10,000): p<0.05 (*), p<0.01 (**), p<0.001(***).

However, when the movement was supported by a 40 Hz subthreshold conditioning train targeting TRI muscles, all the participants enhanced their ability to suppress evoked antagonist activity. With respect to the peak-to-peak amplitude of the reflexes evoked in the BIC at rest, SCS04 showed a reduction of 91%, SCS07 74% and SCS08 90% (Figure 4b, blue bars). We thus directly compared the modulation of the BIC PRM-reflexes when participants performed the movement with and without the support of the stimulation. Similarly to the results found in the rest condition, we observed a clear reduction in the peak-to-peak amplitude of the PRM reflexes evoked in the antagonist muscles (Figure 4c, SCS04 −74%, SCS07 −20%, SCS08 −36% compared to stimulation off). Thus, during active movement, SCS appears to enhance inhibitory effects on antagonistic muscles.

Taken together, the reductions in CMAP amplitude observed during tonic SCS in both the rest and active movement conditions support our hypotheses that SCS engages reciprocal inhibitory circuits in the spinal cord. Importantly, the central regulation of these inhibitory circuits is preserved even after moderate damage to the corticospinal tract and is enhanced with SCS.

### Spinal cord stimulation inhibits firing rates in antagonist motoneurons via postsynaptic inhibition

Although our previous results were consistent with findings suggesting that the inhibitory effects are mediated by the Ia inhibitory interneuron^21,23^, to further confirm the nature of this interaction we analyzed changes in motor unit spiking caused by tonic SCS. We hypothesized that if SCS engages this inhibitory disynaptic connection, then the firing rate of antagonist motoneurons should slow or stop following the SCS pulses. We tested this hypothesis in two participants (SCS07 and SCS08) by using high density electromyography (HDEMG) to detect motor unit spiking in the BIC while the participants produced a sustained elbow flexion torque isometrically for one minute at 30% of their maximum voluntary contraction (MVC) (Figure 5a bottom left panel). The HDEMG signals were measured using a 64-channel array placed on the BIC and decomposed using the convolution kernel compensation method to isolate motor unit action potentials^34^ (Figure 5a top left and right panels). To study the effects of SCS on BIC motoneurons, we applied a conditioning train of tonic stimulation at 40 Hz through the SCS lead contact targeting TRI, similar to the previous conditions. We thus expected that the BIC motoneurons would slow or stop firing following the SCS pulse confirming the interaction through the Ia inhibitory interneuron. Visibly, the peristimulus time histogram (PSTH) and peristimulus frequencygram (PSF)^35^ of the detected BIC motoneurons showed a transient pause in spiking at approximately 15 ms latency (SCS07 average latency 14.3 ms; SCS08 average latency 15.2 ms) from the stimulation pulse (Figure 5b and Extended Data Figure 2a,b). To account for the time needed by the action potential to travel along the motor axons and reach the BIC, we then measured the average latency of responses evoked by suprathreshold stimulation pulses at rest (Extended Data Figure 2c). When accounting for this time delay (SCS07 9.7 ms; SCS08 8.3 ms; Figure 5b and Extended Data Figure 2c), the average inhibition time onset was less than 10 ms (SCS07 4.6 ms and SCS08 6.9 ms) consistent with the timing of disynaptic inhibition ^36,37^. These results therefore support our hypothesis that the effects of SCS are not limited to the monosynaptic connection with α-motoneurons instead it extends to neighboring spinal inhibitory interneurons thus potentiating postsynaptic inhibition.

**Figure 5.**
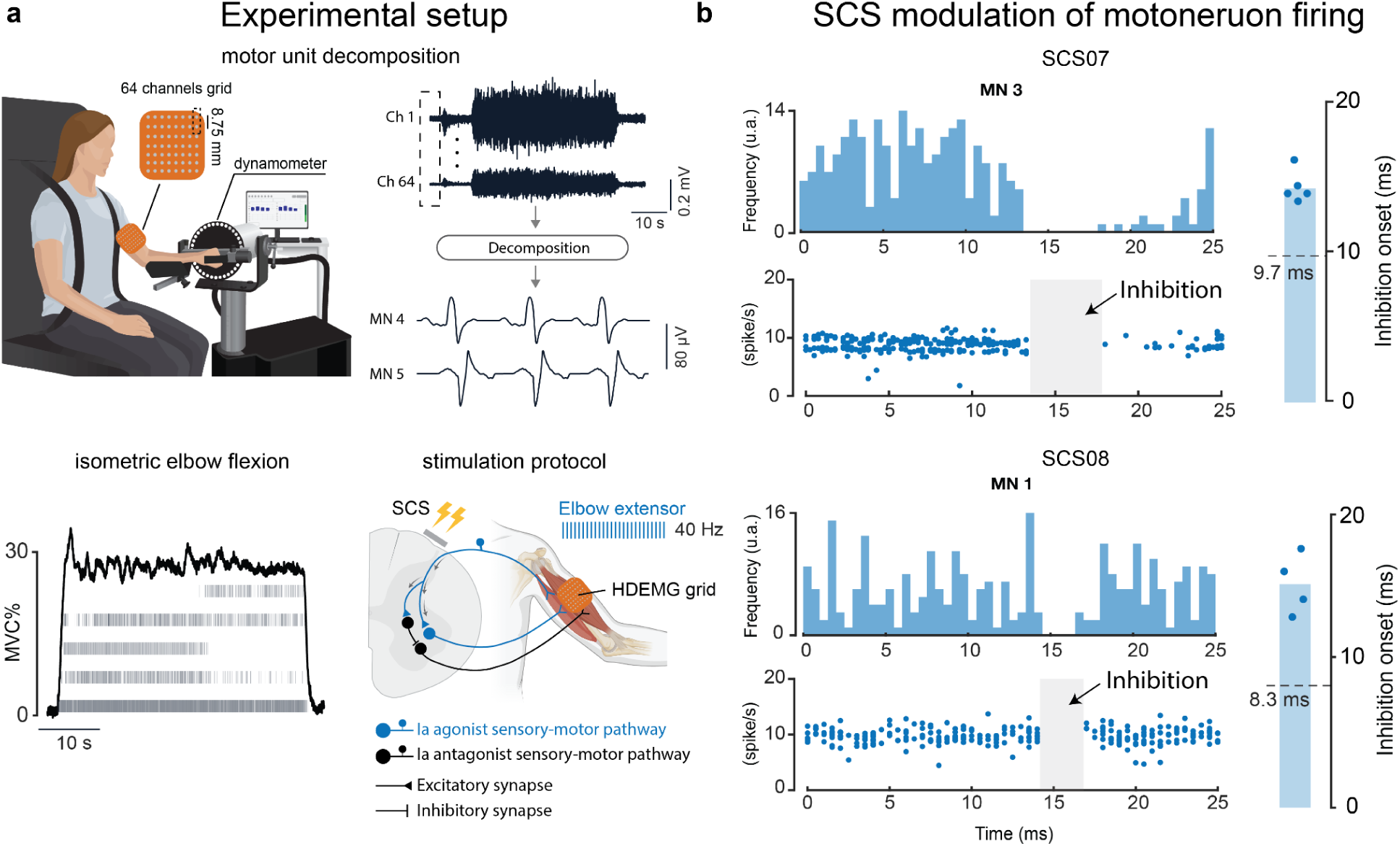
Inhibitory effects of SCS on motoneuron firing: **a**, Top: Description of the experimental setup. Participants were seated in the dynamometer (HUMAC NORM) used to measure the elbow torque produced. We used an 8×8 channel flexible grid placed over the Biceps Brachii to record high density electromyographic activity (HDEMG). The HDEMG recordings were then decomposed into spike trains for individual motoneurons using the convolution kernel compensation (CKC) method^34^. Bottom left: example of torque profile (black) generated by SCS08 during the task overlaid on raster plots of the single motoneurons’ spike trains (gray). Bottom right: simplified schematic of the spinal circuit and stimulation protocol applied with HDEMG grid location. The circuit includes antagonist sensory afferents recruiting α-motoneurons innervating Biceps Brachii (black); agonist sensory afferents recruiting both α-motoneurons innervating Triceps Brachii (TRI) (blue) and Ia inhibitory interneurons innervating α-motoneurons of the Biceps Brachii (BIC) (black). During the task, a 40 Hz stimulation train is applied to target the sensory afferents recruiting TRI. **b,** Top left: Examples of PSTH and PSF for a motor unit in SCS07 and SCS08. Top right: bar plots show the mean value of the inhibition onset delay. The circle markers indicate the time delay between the stimulation pulse and the onset of inhibition for each motor neuron (MN). The dashed line on each plot indicates the average conduction latency of evoked action potentials recorded on the biceps brachii at rest.

## DISCUSSION

In this study, we examined the effects of tonic spinal cord stimulation (SCS) on muscle coordination in three individuals with chronic post-stroke hemiparesis. We found that SCS enhanced reciprocal inhibition between antagonistic muscles, thereby reducing abnormal co-activation and improving the neuromotor control of reaching movements. Our results demonstrate that the modulatory effects of tonic SCS on spinal motoneuron activity are not limited to the well-studied monosynaptic excitation of motoneurons^15–19^, but also extend to inhibitory pathways. Together, these findings provide new insight into the spinal circuits influenced by SCS and support its potential as a neurotechnology to promote motor recovery after stroke.

While recent studies^33^ have focused on how the brain can leverage spinal cord stimulation to enhance motor recovery, here we show that SCS can directly modulate spinal networks responsible for coordinated motor output. We demonstrate that stimulation alone is sufficient to rebalance these fundamental circuits, leading to striking improvements in motor performance after stroke.

A hallmark impairment following stroke is abnormal co-activation of agonist and antagonist muscles^6,38^, often described as pathological synergies and associated with loss of dexterous motor control. These maladaptive patterns restrict range of motion, compromise functional independence, and have been directly linked to corticospinal damage^8,9^. Our study shows that, even in the presence of moderate cortico-spinal damage, SCS can immediately reduce such abnormal co-activation by recruiting spinal inhibitory interneurons. Our findings conceptually align with the work of Mugler et al. (2019)^39^, who developed a myoelectric computer interface (MyoCI) to provide real-time feedback aimed at reducing abnormal co-activation in stroke survivors. In that study, repeated practice with MyoCI promoted motor learning and enabled participants to volitionally decouple muscle pairs, leading to improvements in spasticity, motor impairments and functional outcomes. While our results demonstrate that SCS can achieve similar reductions in co-activation without requiring training, we believe that SCS may complement activity-based rehabilitation methods serving as a substrate upon which training-based interventions can further build, thereby accelerating functional recovery.

It remains unclear whether this abnormal muscle coordination after stroke originates in the brain, due to an imbalanced common drive to the spinal circuits^6^, or in the spinal cord itself, as a consequence of hyperexcitable motoneurons from reduced inhibition^40^. Nevertheless, developing technologies capable of rebalancing sensorimotor networks is crucial for reducing functional limitations and improving quality of life for stroke survivors. Thompson and colleagues^41,42^ have indeed demonstrated that inducing long-term plasticity within spinal inhibitory circuits through operant conditioning of spinal reflexes can result in meaningful motor improvements in people with spinal cord injury. In their studies, H-reflex down-conditioning reduced hyperreflexia, improved gait kinematics, and enhanced walking speed. These results align perfectly with our findings: we show that direct modulation of spinal inhibitory circuits—and specifically the reduction of antagonist muscle excitability—can strongly drive improvements in arm kinematics after stroke. Hence, biasing spinal circuits toward greater inhibition of antagonistic muscles, whether through operant conditioning or electrical stimulation, can restore more normal patterns of motor control after a neural lesion.

At the mechanistic level, the short-latency inhibitory effects observed in our study are consistent with disynaptic reciprocal inhibition mediated by the Ia inhibitory interneurons^21–26^. These interneurons receive input from group Ia afferents of agonist muscles and project to antagonist motoneurons, thereby coordinating reciprocal muscle activity. While our results indicate that SCS targets this pathway to enhance muscle coordination, we cannot exclude the possibility that other Ia-driven circuits—such as central pattern generators, excitatory premotor interneurons or presynaptic inhibitory neurons—are also recruited by stimulation and contribute to the observed improvements in motor control.

### Clinical implications

Reducing abnormal co-activation is a critical therapeutic target in stroke rehabilitation. Current approaches such as botulinum toxin injections, orthotic devices, or intensive task-specific training often provide only partial or transient benefits. By contrast, here we show that SCS could immediately normalize spinal circuitry, which translated into measurable improvements in reaching speed, smoothness, and path efficiency. Notably, even the most impaired participant (SCS04, FM = 23) exhibited substantial motor improvements, suggesting that also individuals with limited residual control may derive particular benefit from this technology.

Although in this study we focused on immediate improvements in upper limb motor control and muscle coordination, the ability of SCS to re-engage inhibitory circuits raises the possibility that it could also mitigate spasticity, another disabling consequence of stroke. For example, Minassian and colleagues^36^ have shown that non-invasive stimulation of proprioceptive fibers can transiently modulate both pre- and post-synaptic inhibition, alleviating spasticity in people with spinal cord injury. Such findings support the idea that restoring deficient inhibitory mechanisms through SCS may benefit other aspects of recovery, not only motor control.

Hence, the clinical potential of SCS extends beyond its immediate effects. By enhancing muscle coordination and potentially reducing spasticity, SCS may promote neuroplasticity and motor learning. This idea aligns with the negotiated equilibrium model of spinal cord function proposed by Wolpaw and colleagues^42,43^, which suggests that targeted modulation of specific spinal pathways can trigger broader adaptive plasticity with benefits across multiple behaviors. In this framework, SCS may act both as a facilitator of immediate motor improvements and as a driver of longer-term reorganization when combined with rehabilitation training.

### Limitations

In this study we designed a battery of tests aimed at measuring the effects of spinal cord stimulation on upper limb motor control and the potential neuromodulation of spinal inhibitory circuits after stroke. While providing support to our results with proper sham conditions, we acknowledge that the limited number of participants (n=3) and the limited range of impairment level restrict the generalizability of our findings. Moreover, multiple studies reported the impact of stimulation parameters to the neuromodulatory effects caused by the stimulation^44,45^. Although we adopted stimulation parameters that were tuned to enhance motor control, it’s worth highlighting that the effect of parameters such as stimulation amplitude and frequency may affect the results^46^. Moreover, unlike the studies of Thomson and colleagues^42^ and Minassian and colleagues^36^, we did not assess the persistence of inhibitory modulation and functional gains after the stimulation was turned off. Thus, further studies are needed to explore these effects over longer timescales, across a wider range of stimulation parameters, and in larger, more diverse cohorts to better establish clinical relevance.

## Conclusion

In this study, we demonstrate that spinal cord stimulation (SCS) engages inhibitory spinal circuits to improve arm motor control in people with chronic stroke. The recruitment of reciprocal inhibitory pathways is particularly important for individuals with chronic hemiparesis, who often struggle with abnormal muscle coordination^6,7^ that restricts movement and reduces quality of life. Our findings also provide mechanistic insight into the neural circuits influenced by SCS and highlight its potential as a therapeutic tool to restore motor function. When combined with training-based interventions, SCS may represent a powerful strategy to promote recovery of upper limb function after stroke.

## METHODS

### Trial information

All experimental protocols were approved by the University of Pittsburgh Institutional Review Board (IRB) (protocol STUDY19090210). Participants were part of an ongoing clinical trial^14^ whose details can be found on ClinicalTrials.gov (NCT04512690). All the participants provided informed consent according to the procedure approved by the IRB of the University of Pittsburgh and participants were compensated for each day of the trial and for travel and lodging during the study period.

### X-ray

X-ray images were acquired at weekly time points in both axial and sagittal views to ensure the stability of lead position.

### Lesion segmentation

Magnetic resonance imaging (MRI) was acquired using a 3T Prisma system (Siemens) using a 64-channel head and neck coil. A T1-weighted structural image was captured using a magnetization-prepared rapid gradient echo sequence (repetition time = 2,300 ms; echo time = 2.9 ms; field of view = 256 × 256 mm2; 192 slices, slice thickness = 1.0 mm, in-plane resolution = 1.0 × 1.0 mm). Lesion segmentation was performed manually for each slice of the sequence using the MRIcron image viewer (NITRC); the resulting region of interest was smoothed on all planes using a Gaussian smoothing kernel with full-width at a half-maximum of 2 mm. MRIcroGL (NITRC) was used to visualize and export the resulting segmented overlays.

### Spinal cord stimulation system

The system used to deliver the spinal cord stimulation was the same as the one described in previous studies^14^. Briefly, two 8-contact linear SCS leads were implanted in the lateral aspect of the cervical spinal cord to target the dorsal roots innervating arm and hand muscles^14,18^(C3 to T1 spinal segments). Stimulation pulses were delivered using a biphasic constant current stimulator (DS8R; Digitimer, Welwyn, Garden City, England) connected to a 1-to-8 channel multiplexer (D188, Digitimer, Welwyn, Garden City, England). Stimulation amplitude and frequency was then controlled using a custom-made Matlab based (R2021 a/b, The Mathworks Inc., Natick, MA) Graphical User Interface (GUI). Participants took part in the clinical trial for 29 days after which the SCS leads were removed.

### Clinical evaluations

#### Fugl-Meyer upper extremity assessment

The Fugl-Meyer upper extremity assessment is a standard measure of upper-limb motor control and sensory function^47^. In total there are 126 possible points divided among seven categories, however, in this manuscript we reported only the motor and sensory subscores which have a maximum of 66 and 12 points, respectively. The same trained therapist administered the exam at four different time points: prestudy; mid-study (∼ two weeks after implant); end of study (four weeks); and post-study (∼ one month after implant removal). In the manuscript we reported only the pre-study scores.

### Kinematics and EMG analysis

#### Planar reaching task

To isolate the kinematics analysis to the elbow joint we used a robotic augmented reality exoskeleton system (KINARM, Kingston, ON, Canada). Participants were secured in a wheelchair with adjustable height and limb supports to avoid the contribution of gravity. The task was then displayed onto a dichroic augmented reality display in front of the individuals. Specifically, participants were asked to reach one of the three virtual targets while receiving the feedback of the hand position (Figure 2a). On each trial, the task started with the robot moving the participant’s arm to the starting position. To signal the beginning of the movement, an audio cue was played after a randomized time delay of 100-700 ms. The reaching phase was considered completed when the participant kept the index finger for 500 ms within a 0.5 cm radius respect to the target. If participants were unable to reach the target within 10 seconds from the audio cue, the robot moved the arm back to the starting position and the next target was presented. Target position was adjusted with respect to each participant’s range of motion to avoid patient discomfort while performing the task. In the manuscript we reported the results of n = 12 trials for both stimulation Off and On, n = 20 trials for the Sham stimulation in SCS04; n=30 trials for both stimulation Off and On and n= 9 trials for the sham stimulation in SCS07; n= 9 trials for stimulation Off, On and Sham stimulation for SCS08.

#### Kinematics performance metrics

To quantify movement performance we measured three kinematics metrics for each trial: time to reach the target (s), number of elbow speed peaks (u.a.) and path efficiency (u.a.). The time to reach the target was reported in seconds and measured as the time elapsed from the starting audio cue to the completion of the reaching phase. A shorter time indicates faster movements. The smoothness of the movement was quantified as the number of peaks in the elbow speed profile during the same time interval. Fewer peaks indicate smoother movements. The Matlab function *findpeaks* with minimum peak height of 0.1 was used to identify the peaks in the speed profile. Finally, the path efficiency was measured as the Euclidean distance between the starting position and the end target normalized with respect to the total hand path length within the time interval describing the reaching phase. The equation that describes the quantification of this metric is the following: 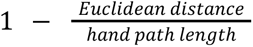. A value closer to 0 therefore indicates trajectories closer to a straight line. Data processing and analysis was done with Maltab R2023b (The Mathworks Inc., Natick, MA).

#### Electromyographic activity

EMG data from the upper limb muscles were acquired using the wireless Avanti sensors (Trigno, Delsys Inc.) and synchronized with the kinematics data using a data acquisition (DAQ) system with sampling frequency of 8000 Hz. For the quantification of the muscle activity we reported only the main muscles involved in the planar reaching movement (Elbow extensors: SCS04,SCS07 and SCS08 Triceps Brachii; Elbow flexors: SCS04,SCS07 Brachioradialis and SCS08 Biceps Brachii). EMG data were band-pass filtered (20 – 500 Hz, 3th order Butterworth digital filter), full-wave rectified and smoothed with moving average sliding window of 500 ms. We then measured the mean EMG value of the filtered signals for each of the kinematics trials. To account for possible stimulation artifacts, we visually inspected the raw EMG traces and blanked with a 10 ms window centered around the stimulation pulse when we detected the presence of artifacts. Data processing and analysis was done with Maltab R2023b (The Mathworks Inc., Natick, MA).

### Recruitment curves and stimulation protocols

#### Recruitment curves

To evaluate the specificity of SCS in recruiting individual motor pools and account for accurate leads placement, recruitment curves were performed for all the SCS leads contact on the first day of testing after the implant (Figure 2a bottom panels and Extended Data Figure 1a). We used the wireless EMG system (Trigno, Delsys Inc.) to record the compound action potentials (CMAPs) elicited by SCS pulses in the upper limb muscles. The SCS pulses were delivered at 1 Hz while gradually increasing the stimulation amplitude. We then measured the peak-to-peak amplitude of the evoked muscles responses and normalized with respect to the average value of the three highest amplitudes recorded on that muscle across all the trials. The peak-to-peak value was measured as the max-min difference within a 40 ms window after the SCS pulse. To avoid possible stimulation artifacts we blanked the first 6 ms following the stimulus.

#### Stimulation parameters

Based on the recruitment curves we selected the combinations of SCS lead contacts and amplitude which best targeted elbow flexors (i.e., Sham) and extensors (i.e., stimulation On). Specifically the stimulation parameters were: SCS04, 8R 60 Hz 4.4 mA (On) - 1C 60 Hz 1.9 mA (Sham); SCS07, 4R 40 Hz 1.4 mA (On) - 1R 40 Hz 1.4 mA (Sham); SCS08, 7R 60 Hz 2.6 mA (On) - 2R 60 Hz 3.2 mA (Sham).

### Posterior root-muscle reflexes analysis

To assess the modulation of the posterior root-muscle (PRM) reflexes across the conditions (Figure 3 and Figure 4) we measured the peak-to-peak amplitude of the compound action potentials evoked in the Biceps Brachii (i.e., elbow flexor) of all the participants. We first identified the SCS lead contact and stimulation amplitude which evoked consistent responses in the target muscle at rest. Thus we fixed the stimulation parameters for both the condition at rest (Figure 3) and during movement (Figure 4). We then applied the conditioning stimulation trains through the contact recruiting elbow extensors (i.e., stimulation On) and forearm/hand muscles (i.e., Sham). Specifically the stimulation parameters for each participant and condition were the following.

#### Rest condition (Figure 3)

SCS04, Off : 5R 2 Hz 4.0 mA (Test pulse); On: 5R 2 Hz 4.0 mA (Test pulse) + 8R 40 Hz 3.8 mA (Conditioning train); Sham: 5R 2 Hz 4.0 mA (Test pulse) + 5C 40 Hz 2.4 mA (Conditioning train). SCS07, off : 1R 2 Hz 1.4 mA (Test pulse); On: 1R 2 Hz 1.4 mA (Test pulse) + 6R 40 Hz 1.4 mA (Conditioning train); Sham: 1R 2 Hz 1.4 mA (Test pulse) + 6C 40 Hz 1.4 mA (Conditioning train). SCS08, Off : 4R 2 Hz 3.0 mA (Test pulse); On: 4R 2 Hz 3.0 mA (Test pulse) + 6R 40 Hz 3.0 mA (Conditioning train); Sham: 4R 2 Hz 3.0 mA (Test pulse) + 5C 60 Hz 3.0 mA (Conditioning train).

#### Movement condition (Figure 4)

SCS04, Off : 5R 2 Hz 4.0 mA (Test pulse); On: 5R 2 Hz 4.0 mA (Test pulse) + 8R 40 Hz 3.8 mA (Conditioning train); SCS07, Off : 1R 2 Hz 2.2 mA (Test pulse); On: 1R 2 Hz 2.2 mA (Test pulse) + 6R 40 Hz 1.4 mA (Conditioning train). SCS08, Off : 4R 2 Hz 3.4 mA (Test pulse); On: 4R 2 Hz 3.4 mA (Test pulse) + 6R 40 Hz 3.0 mA (Conditioning train).

In the condition at rest (Figure 3) the participants were sitting in the KINARM exoskeleton to support the weight of the paretic limb against gravity. A member of the study team was then holding the arm in a fully extended position for the entire duration of the test. Participants were asked to relax, neither to resist, follow, nor facilitate the movements. When the stimulation protocol was applied during movement, instead, participants were instructed to reach only one of the three virtual targets used for the planar reaching task. The peak-to-peak amplitude of the evoked potentials was then measured as the max-min difference within a 40 ms window after the SCS pulse targeting the Biceps Brachii. To avoid possible stimulation artifacts we blanked the first 6 ms following the stimulus. We report here the number (n) of collected responses for each participant and condition. *Rest condition* (Figure 3). SCS04: stim Off (baseline), n = 24; stim On, n = 24; Sham stim, n = 24. SCS07: stim Off (baseline), n = 54; stim on, n = 54; sham stim, n = 54. SCS08: stim off (baseline), n = 54; stim On, n = 54; Sham stim n = 54. *Movement condition* (Figure 4). SCS04: stim Off, n = 24; stim On, n = 24. SCS07: stim Off, n = 30; stim On, n = 30. SCS08: stim Off, n = 30; stim On, n = 30. To compare the conditions we then used bootstrap to compute the 95% confidence interval (CI) of the peak-to-peak amplitudes (N=10000). The middle circle in the box plots shows the mean peak-to-peak value. Error bars show the 95% CI.

### Single motoneuron firing rate analysis

#### Motor unit decomposition

We recorded high density surface electromyography (HDEMG) from the Biceps Brachii while two participants (SCS07 and SCS08) performed isometric elbow flexion for 1 minute sustaining 30% of their maximum voluntary contraction (MVC). HDEMGs signals were not recorded for SCS04 therefore he was excluded from the analysis. The elbow torque was measured with a robotic platform (HUMAC NORM, CSMi) and provided in real time with a custom-made Python-based graphical user interface. An 8×8 channel flexible HDEMG grid electrode with 8.75 mm distance between electrodes was used for the recordings. Conductive gel was used to reduce the skin-electrode impedance. EMG recordings were acquired in monopolar configuration using the TMSi Saga 64+ high density amplifier at a sampling rate of 2 kHz with the reference electrode placed over the lateral epicondyle of the elbow. We then decompose the HDEMG recordings into individual motoneurons spike trains using DEMUSE tool software which exploits the convolution kernel compensation (CKC) method^34^. The results of this automatic decomposition were manually edited following the standard procedure already described ^48,49^. Only motoneurons with pulse-to-noise ratio ≥ 20 dB were selected for the analysis.

#### Inhibited motoneuron identification

Based on our hypothesis, if the stimulation of the sensory afferent innervating the Triceps Brachii enhances the activity of the Ia inhibitory interneurons innervating the Biceps Brachii, then the detected motoneurons should stop firing after a certain delay from the SCS pulses. Thus, we plotted the peristimulus time histogram (PSTH) (Figure 5b) and the peristimulus frequencygram (PSF) (Figure 5b and Extended Data Figure 2a,b) around the SCS pulses. For the PSTH we quantified the number of spikes occurring in bins of 0.5 ms. The PSF instead, represents the instantaneous discharge rate of each motoneuron at a certain delay from the stimulation pulse^35^. Therefore, when the motoneurons innervating the Biceps Brachii were inhibited by the stimulation pulse, no spikes could be detected in the PSTH and PSF after a certain delay. To quantify the inhibition time onset, we first manually measured the time delay after which we could detect the absence of spikes on the PSF of each detected motoneuron. Motoneurons which showed an inhibition time onset lower than 3 ms were excluded from the estimate of the average inhibition latency given that they might be the result of longer latency inhibition^23^ and therefore erroneously bias the results. However, only motoneuron number 2 for SCS07 was excluded for this reason (Extended Data Figure 2a). Motoneurons which did not show absence of spikes following the stimulation pulse were also excluded from the estimate of the inhibition latency. Only motoneuron number 5 for SCS08 was excluded for this reason (Extended Data Figure 2b). Then, to account for the time needed by the action potential to travel along the motor axon, we measured the latencies of the PRM-reflexes evoked in the Biceps Brachii muscle at rest while delivering supra-threshold SCS pulses. The latencies were measured manually and reported as the milliseconds between onset of the stimulus and first deflection of evoked EMG responses (Extended Data Figure 2c). We thus subtracted the average latency value of the action potential from the measured time delays after which no spikes were detected to obtain the inhibition time onset following the SCS pulses.

### Statistics

In this manuscript we assessed the statistical significance in the comparison of the stimulation On and stimulation Off conditions ( Off vs On; Off vs Sham). All the statistical comparisons of the means were performed using the bootstrap method. This method is a non parametric approach which makes no assumptions about the distribution of the observed data. Instead, it uses resampling to construct empirical confidence intervals (CIs) of the quantities of interest. For each comparison, we constructed bootstrap samples by drawing a sample with replacement from the observed measurements while preserving the sample size. Thus, we constructed N = 10000 bootstrapped samples for each measurement and calculated the difference in means of the resampled data. Of the resulting values, we then computed the 95% CI by identifying the 2.5th and 97.5th quantile. The null hypothesis of no difference in the mean was rejected if 0 was not included in the 95% CI. To compute the p-value, we centered the resulting bootstrap distribution of the difference between the means at 0 (i.e., null hypothesis). We then computed the p-value as the probability of observing a value either equal to the difference in means of the quantities of interest or more extreme under the null hypothesis.

## Data Availability

All data produced in the present study are available upon reasonable request to the authors

## AUTHOR CONTRIBUTIONS

L.B., D.J.W. and M.C. designed the experiments. L.B., N.V., E.S. designed and implemented the stimulation control system, and the hardware and software framework for the experiments. E.C. and L.B. designed and implemented the KINARM motor tasks. S.E. and E.P. designed and performed the MRI-based analyses. A.B. and G.F.W. implemented patient recruitment, eligibility and monitoring and coordinated management of the study. M.C., L.E.F. and P.C.G. designed the neurosurgical approach. P.C.G. and D.P.F implemented and evaluated the neurosurgical procedures and participants’ clinical management protocols. L.B, E.S., N.V., E.C. and R.F. performed the experiments. L.B. created the figures and analyzed the data. L.B., D.J.W., wrote the paper and all authors contributed to its editing.

## ACKNOWLEDGMENTS

The study was executed through the support of National Institutes of Health Brain Initiative grant no. UG3NS123135-01A1 to M.C. and D.J.W. and internal funding from the Department of Neurological Surgery at the University of Pittsburgh to M.C., the Department of Mechanical Engineering and the Neuroscience Institute at Carnegie Mellon University to D.J.W. and the Department of Physical Medicine and Rehabilitation at the University of Pittsburgh to E.P.

## Supplemental information

**Extended data figure 1.**
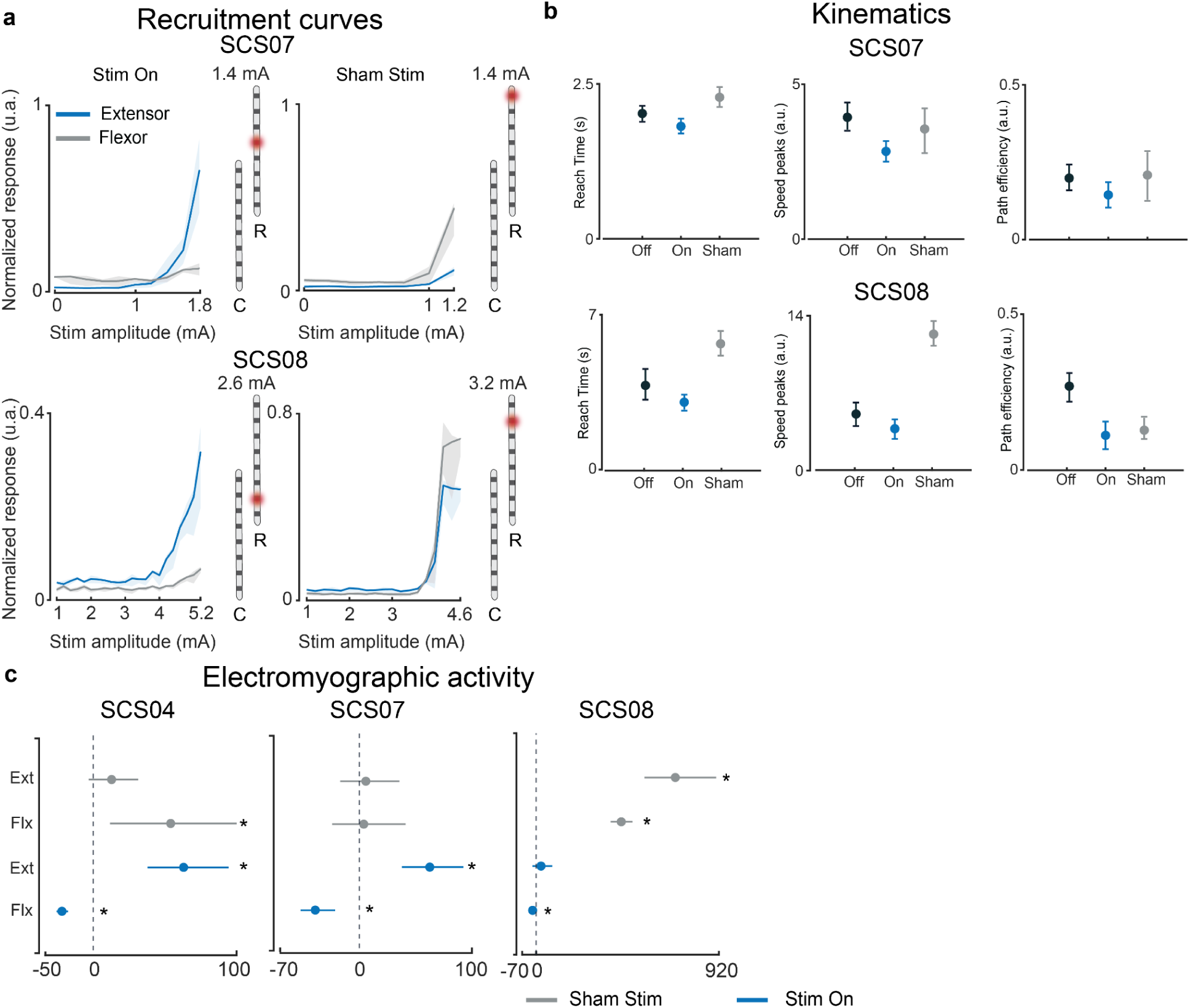
Planar reaching kinematics and muscle activity: **a**, Top: recruitment curves for contact 4R (left) and contact 1R (right) and normalized evoked responses for triceps brachii (blue, elbow extensor) and biceps brachii (gray, elbow flexor). Bottom: recruitment curves for contact 7R (left) and contact 2R (right) and normalized evoked responses for triceps brachii (blue, elbow extensor) and biceps brachii (gray, elbow flexor). **b**,Quantification of the mean values of the kinematics metrics used to assess the effect of SCS (On) and Sham stimulation with respect to stimulation Off for SCS07 (top) and SCS08 (bottom). **c**, Quantification of percent change of the muscles EMG activity with respect to stimulation Off (EXT: triceps brachii with stimulation On (blue) and Sham (gray) stimulation; FLX: brachioradialis (SCS04, SCS07), biceps brachii (SCS08) with agonist (blue) and Sham (gray) stimulation). The middle circle on plots indicates the mean value of the measurements. All error bars indicate the 95% confidence interval computed with bootstrap N=10,000. The single asterisk indicates statistical significance and rejection of the null hypothesis of no difference with a 95% CI.

**Extended data figure 2.**
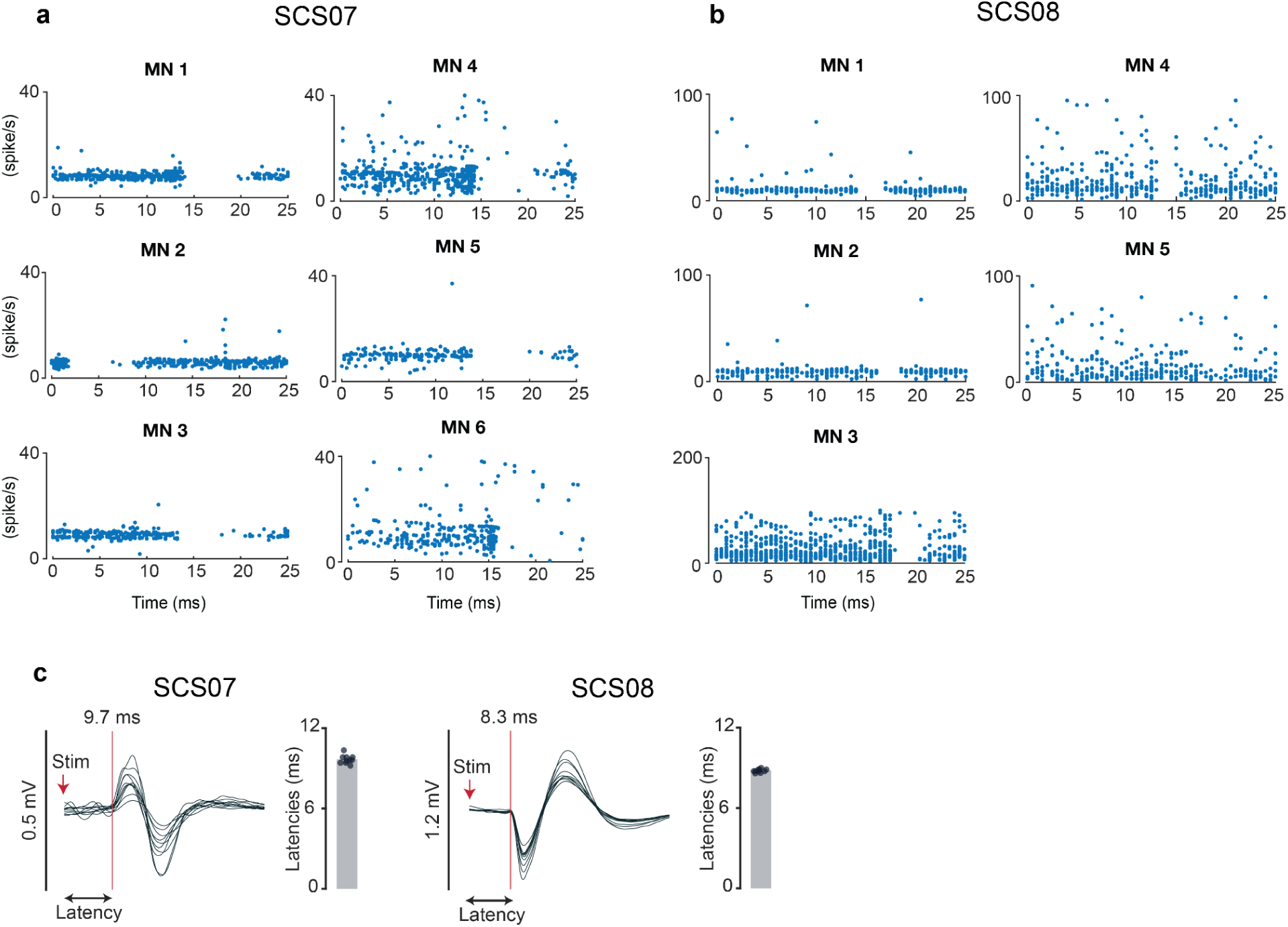
Peristimulus frequencygram (PSF) and action potential latencies: **a**,**b** Peristimulus frequencygram of all the detected motoneurons (MNs) for SCS07 and SCS08, respectively. Circles on plots indicate the instantaneous firing rate of the MNs at the reported delay from the SCS pulses. The interval considered spans 0 ms to 25 ms post-stimulus, which is the stimulation time period for the 40 Hz tonic SCS used in this experiment. **c**, Latencies of the PRM-reflexes evoked in the biceps brachii as an estimate of the action potential propagation delay for SCS07 and SCS08. Bar plots show the mean value of the action potential delay with respect to the SCS pulses. Circles on plots indicate the time delay between the stimulation pulse and the recorded action potential for n = 10 evoked responses. Evoked responses were recorded at rest while delivering supra-threshold SCS targeting Ia sensory afferents innervating Biceps Brachii.

## Notes

### Competing Interest Statement

M.C. and D.J.W. hold patents in relation to spinal cord stimulation. M.C., D.J.W., and P.C.G. are founders and board members of Reach Neuro, a company developing spinal cord stimulation technologies for stroke. E.P. has interests in Reach Neuro because of marital status with M.C.

### Clinical Trial

NCT04512690

### Author Declarations

University of Pittsburgh Institutional Review Board (IRB) (protocol STUDY19090210) gave ethical approval for this work

